# Genomic descriptive study of human Parvovirus B19 circulation during 2024/2025 in the State of Rio de Janeiro, Brazil

**DOI:** 10.64898/2026.07.03.26357244

**Authors:** Melise Chaves Silveira, Renata Campos Azevedo, Alessandra Pavan Lamarca, Maria Angélica Arpon Marandino Guimarães, Claudia Maria Braga de Mello, Adriana Cardoso Camargo, Alexandra Lehmkuhl Gerber, Ana Paula C. Guimarães, Andréa Cony Cavalcanti, Ana Tereza Ribeiro Vasconcelos

## Abstract

**Purpose:** Human Parvovirus B19 (B19V) infection is associated with a broad spectrum of clinical manifestations, including erythema infectiosum, arthropathy, transient red cell aplasia, hepatitis, and adverse fetal outcomes. Due to nonspecific presentations and limited routine testing, B19V infection often remains undiagnosed. Despite recent reports of increased B19V activity worldwide, contemporary data on its presence in Brazil remain scarce. We investigated the circulation of underdiagnosed pathogens in patients with suspected infectious diseases, prioritizing severe and fatal cases.

**Methods:** In this descriptive study, hybrid capture-based whole-genome sequencing was used for a broad-range viral detection and characterization. We analyzed 472 clinical specimens selected for diagnostic investigation between 2024 and 2025, according to surveillance criteria for respiratory and arboviral infections, referred to the Public Health Laboratory of the state of Rio de Janeiro, Brazil.

**Results:** B19V was the second most frequently detected viral pathogens, being identified in 190/472 specimens. Of these, just one sample had been previously tested for this pathogen. Thirty-one cases had higher B19V genomic coverage and were therefore selected for further analysis. Notably, B19V was the only virus detected with substantial genomic coverage in nine individuals, including elderly patients, and meningitis cases with B19V-positive cerebrospinal fluid. Phylogenetic analysis showed that recovered genomes clustered within genotype 1A2.

**Conclusions:** B19V was frequently detected in patients undergoing investigation for infectious diseases and its circulation may represent an important and underrecognized pathogen. These findings support the incorporation of B19V testing into diagnostic algorithms for unexplained infectious diseases, especially in patients presenting atypical symptoms.

## Introduction

Parvoviruses are single-stranded DNA viruses infecting a wide range of hosts, from crustaceans to primates. Human Parvovirus B19 (*Primate erythroparvovirus 1*, B19V), a member of the genus *Erythroparvovirus*, is primarily transmitted by the respiratory route, although parenteral transmission occurs [1]. Following the onset of the acute infection, viral DNA may persist in blood, saliva, and respiratory secretions for up to one week [2]. B19V causes diverse manifestations, including erythema infectiosum, polyarthropathy, hematological disorders, and chronic inflammatory conditions. It can also pass through the placenta, causing fetal complications such as hydrops fetalis, congenital anemia, and intrauterine death [3–6].

Clinical diagnosis of B19V infection in the tropics is difficult because its manifestations overlap with those of endemic arboviruses and other rash-associated illnesses [7]. Laboratory diagnosis is conducted via detecting B19V-specific antibodies, although IgM titers may be undetectable during acute infection or in immunocompromised individuals [2,4,8]. Molecular assays are highly sensitive but cannot distinguish infectious virions from naked viral DNA [2]. Despite available commercial assays, standardized diagnostic kits remain limited. Coinfections with B19V are poorly characterized, although associations with hepatitis B virus, hepatitis C virus, cytomegalovirus, and BK polyomavirus have been reported [9].

Despite its clinical importance, B19V remains a neglected pathogen in Brazil, where it is not a notifiable disease and regional incidence estimates are scarce [4]. In the state of Rio de Janeiro, contemporary data is limited to studies focusing on specific clinical syndromes [10,11], with no description of current circulation, genotypic diversity, or the burden of infection in severe and fatal cases. The international resurgence of the B19V post-2019 pandemic [3,8,12–15] underscores the need for updated understanding of the dynamics of the pathogen in the state.

Next-generation sequencing (NGS) has emerged as a promising tool for diagnosing infectious diseases, expanding local screening assays [16]. Capture-based enrichment enables analysis of genomes directly from clinical specimens and has demonstrated high sensitivity for viral targets, even at low viral loads [17,18]. Genomic approaches improve the identification of emerging pathogens and accelerate outbreak response. They may improve B19V laboratory detection, which remains limited and mild clinical manifestations are frequently overlooked [19].

This study used a highly sensitive, broad-range genomic approach to screen for underdiagnosed viral infections in a heterogeneous group of patients attending hospital units in the state of Rio de Janeiro between 2024 and 2025, with particular emphasis on B19V infections. It follows a recent international effort to raise awareness of B19V and may contribute to public health preparedness in Brazil.

## Methods

### Study design and sample selection

This descriptive, observational genomic surveillance study used a convenience sample of clinical specimens collected from patients with suspected infectious diseases, including respiratory and systemic manifestations, in the state of Rio de Janeiro between 2024 and 2025 (Supplementary Table 1). Specimens were obtained through routine public health surveillance activities and referred to the Bioinformatics Laboratory (LABINFO) for whole-genome sequencing (WGS) to characterize the viruses. Sample inclusion was based on referral criteria established by the surveillance system for respiratory and arbovirus infection, with prioritization of severely ill and fatal cases. In total, 472 human samples from various clinical specimens were selected (Supplementary Table 1).

Demographic and clinical data were obtained from laboratory records and, when available, from associated health information systems. Clinical presentations were categorized by referring diagnosis (e.g., respiratory infection, meningitis, or other suspected viral syndromes). Fatal outcomes were recorded when available. Co-detection was defined as the identification of more than one viral agent in a single sample based on genomic sequencing. Due to the study’s retrospective design, the completeness of clinical data varied across samples.

This work was previously approved by the Ethics Committee of Rio de Janeiro State Health Department, number: 7.686.303 and by the Ethics Committee of the Federal University of Rio de Janeiro, number: 4.546.307.

### Sample preparation and whole genome sequencing

Total nucleic acid extraction was performed using the MagMAX Viral/Pathogen Nucleic Acid Isolation Kit (Thermo Fisher Scientific, USA), according to the manufacturer’s instructions. WGS library preparation used the Viral Surveillance Panel v2 (Illumina, USA). This protocol includes pre-enrichment and enrichment steps based on a hybrid-capture approach, incorporating probes targeting more than 200 RNA and DNA viral genomes. Library size and integrity were determined using the TapeStation 4200 System (Agilent Technologies, USA) according to the manufacturer’s instructions. DNA concentration was measured using Qubit 4.0 Fluorometer (Thermo Fisher Scientific, USA). Sequencing was conducted on a NextSeq 2000 platform (Illumina, USA) in paired-end mode, generating 150 bp reads.

### Sequence Data Processing

FASTQ files were examined for quality control using FastQC (https://www.bioinformatics.babraham.ac.uk/projects/fastqc/). Reads were then processed with fastp using default parameters [20]. Alignment of high-quality reads and indexing of the human reference genome (GCA_000001405.29) was performed using Bowtie2 v2.2.6 [21]. SAMTools v1.17 was used to convert SAM to BAM files and to generate FASTQ files of unmapped reads [22].

### Identification of viral genomes

Viral reference genomes were downloaded using accession numbers recovered from VSPv2 v2.7.0 panel summary, (https://help.idm.illumina.com/dragen-microbial-enrichment-plus/dragen-microbial-enrichment-plus, accessed on September 5, 2025). The genome sequences of Rhinovirus A strain RV-A21 and Human rhinovirus NAT001 were added to the reference database (MF043119.1 and NC_038878.1), expanding rhinovirus identification. Unmapped reads from the human genome were used for viral identification. Reads were aligned to the target viral sequences using BWA (version 0.7.17) [23]. SAMTools v1.17 was used to obtain BAM files and to calculate coverage, mean depth and number of reads mapped [22]. BcfTools v1.22 was applied to generate the variants file [24], which was then inputted to BedTools2 v2.31.1 to generate a consensus genome sequence, also considering uncoverage regions [25]. Heatmaps were generated in R using the pheatmap package, with hierarchical clustering based on Euclidean distance and average linkage for both rows and columns [26]. Coverage was used as a marker of viral detection, and the top five coverage values per sample were analyzed. For specific pathogens that share a substantial genome identity, the one with the highest coverage is represented in the heatmap (Human Respiratory Sinusitis Virus A and B; Dengue Virus 1, 2, 3 and 4; Adenovirus A, B, C, D and E; Influenza virus A-H1N1 and A-H3N1; and Herpes simplex virus HSV-1 and HSV-2). Reads per million (RPM) were calculated by dividing the number of reads mapped to the B19V reference genome (NC_000883) by the total number of reads generated, and multiplying the result by 10^6^.

### Phylogenetic construction

Global reference sequences were retrieved from GenBank. Sequences were aligned using the MAFFT v7.522 with the “–auto” option [27]. Maximum-likelihood phylogenetic trees were inferred with IQ-TREE v2.2.2.6, and the substitution model was selected with ModelFinder [28]. Node support was calculated with 1000 ultrafast bootstrap and 1000 SH-aLRT replicates. Bootstrap values ≥ 70% were considered statistically significant and are shown for the corresponding nodes. For the NP1 tree, subsequences were extracted using SeqKit [29] for samples that showed 100% coverage of the *ns1* gene (NC_000883.2:616-2631). Trees were rooted by their midpoint and visualised in iTOL v7.5 (https://itol.embl.de/).

## Results

The viral WGS panel identified B19V as one of the most frequently detected viruses. A total of 190 samples (40.25%) showed some degree of genome coverage relative to the reference sequence. Among all samples, one had undergone prior routine serological testing for B19V, and the result was positive (VSP00003).

For B19V genomes detected in each sample, sequencing metrics and sample source were analyzed (Figure 1). The combined plot showed that genomes with >80% coverage were associated with a mean depth greater than 20x. RPM values above 2.35 (log10) were detected in cerebrospinal fluid (CSF), nasopharyngeal swabs, and serum. A single sample reached 100% reference genome coverage and showed a high number of reads mapped to the B19V reference (VSP00003).

**Figure 1:**
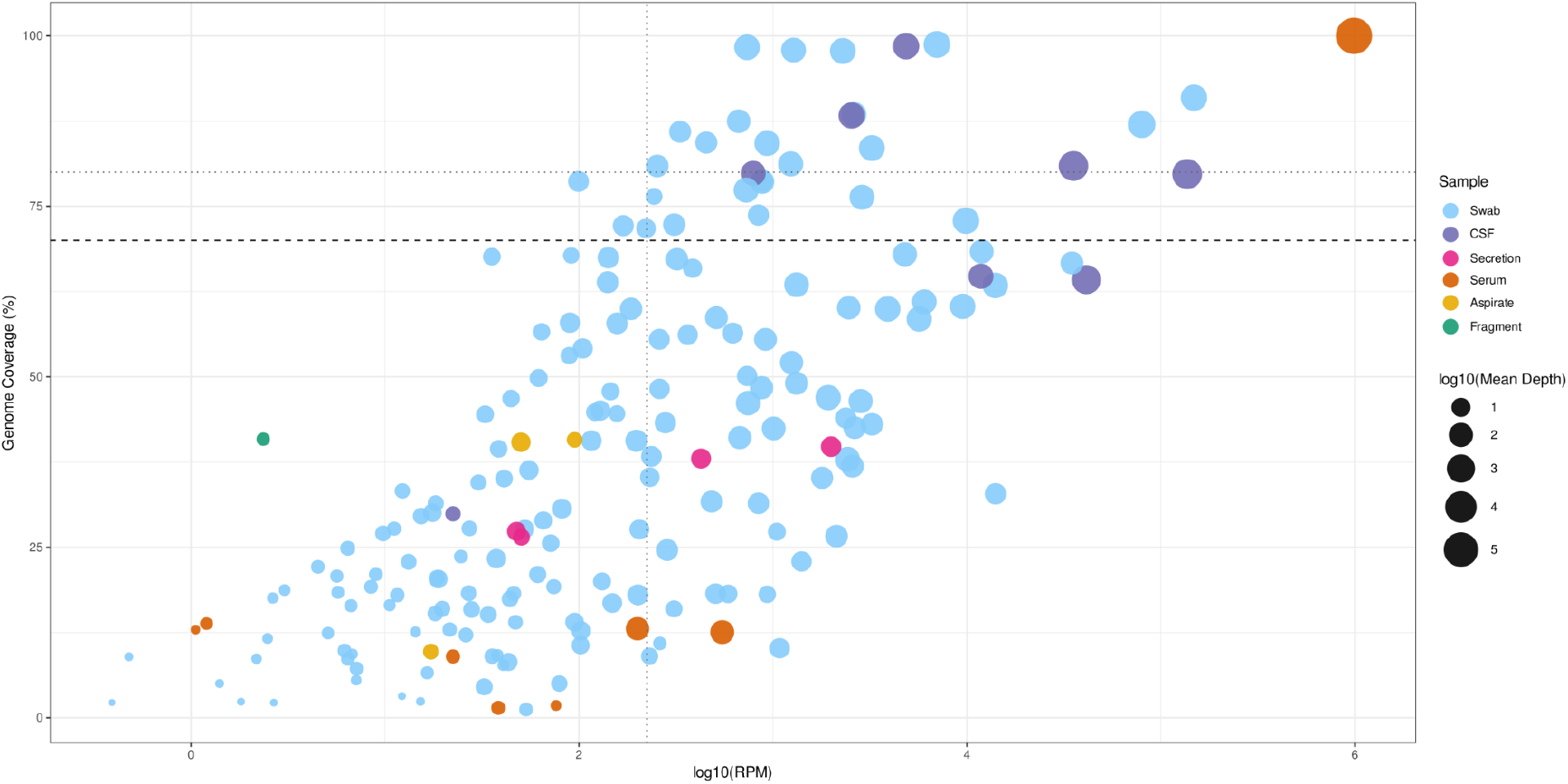
Scatter plot of log_10_ RPM versus B19V genomic coverage (%) for 190 samples. Each point represents a sample, annotated by origin and mean depth of coverage (log10). Point size indicates mean depth of coverage (log10), while color reflects sample origin. The dashed line indicates 70% coverage. Dotted lines indicate coverage above 80% and RPM above 2.35 (log10).

To construct a broader genomic surveillance panel, the genomic coverage threshold was set at 70%, resulting in 31 samples that were carefully analyzed (Figure 2). Overall, this group includes 22 males and 9 females; samples were collected between January 2025 and May 2025 from nasopharyngeal swabs (25), CSF (5), and serum (1). The clinical diagnosis suggested suspected infections by influenza or other respiratory virus (14), COVID-19 (11), meningitis (4), chikungunya virus (1), and HSV (1). For these samples, the heatmap shows that in addition to B19V, other viruses were co-detected, with human respiratory syncytial virus A and B (HRSV-A and HRSV-B) being the most frequently co-occurring.

**Figure 2:**
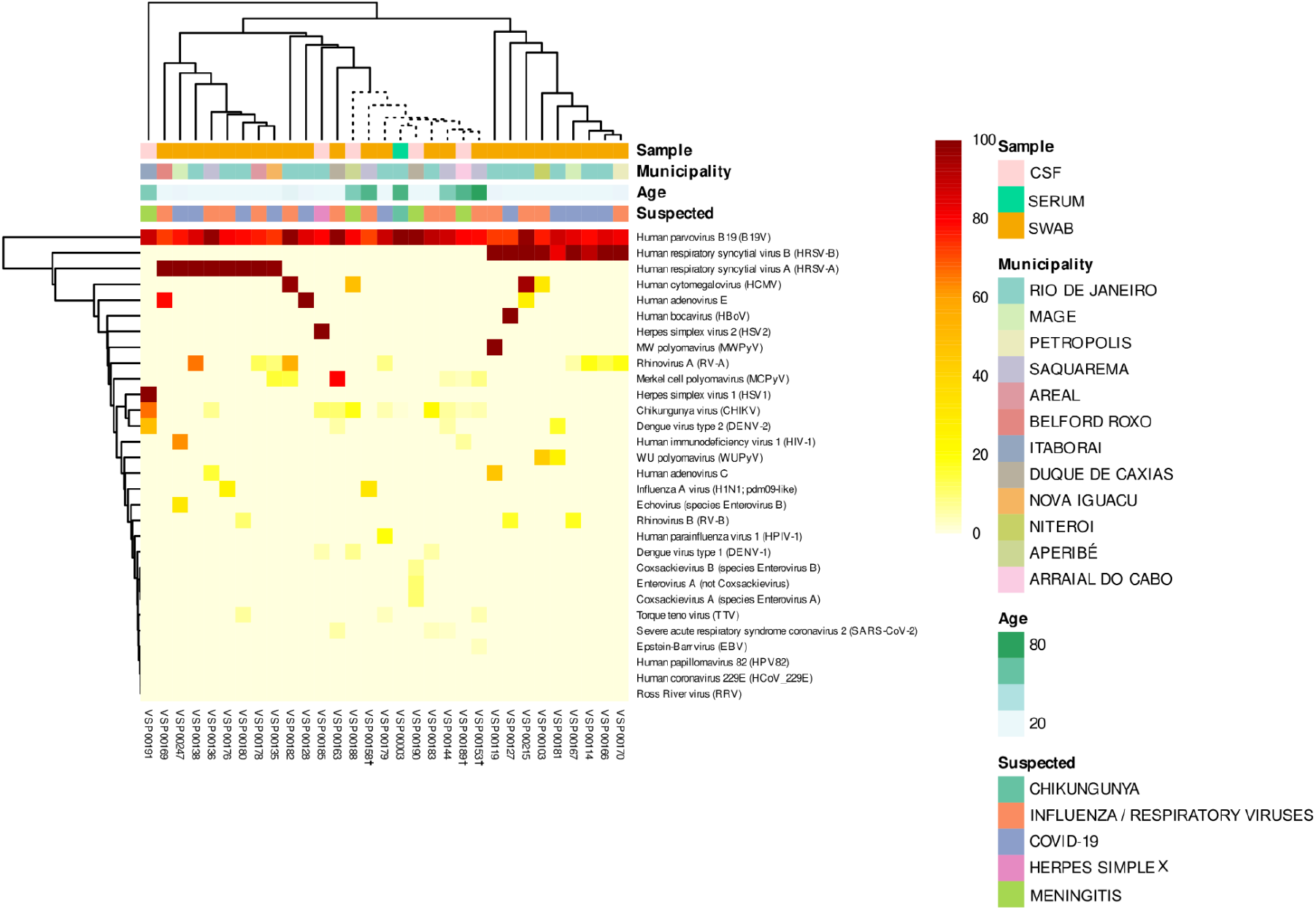
Heatmap displaying genomic coverage of the five most frequently detected viruses across samples above 70% genomic coverage for B19V. Rows represent viruses and columns represent individual samples. Color intensity indicates the percentage of genomic coverage for each virus–sample combination. Sample metadata, including origin, municipality, patient age, and suspected diagnosis, are displayed as annotations. The dotted cluster highlights samples in which B19V was the only virus detected with substantial coverage. Samples from fatal cases are indicated with crosses.

In 9 samples (Figure 2, dotted cluster), B19V was the only virus detected with substantial coverage, including 3 suspected meningitis cases. Among these 9 samples, 4 were obtained from older individuals (≥60) and comprise serum, nasopharyngeal swabs and CSF. All fatal cases (3) occurred within this older age group and were located in the coastal region (Saquarema and Araial do Cabo).

Deaths were registered for 3 of the 31 samples with B19V coverage higher than 70% (Table 1), all from elderly patients. One patient was HIV-1 positive. The co-occurring viruses showed genome coverage below 33% (Supplementary Table 1). The cause of death for one patient was attributed to acute renal failure.

**Table 1:**
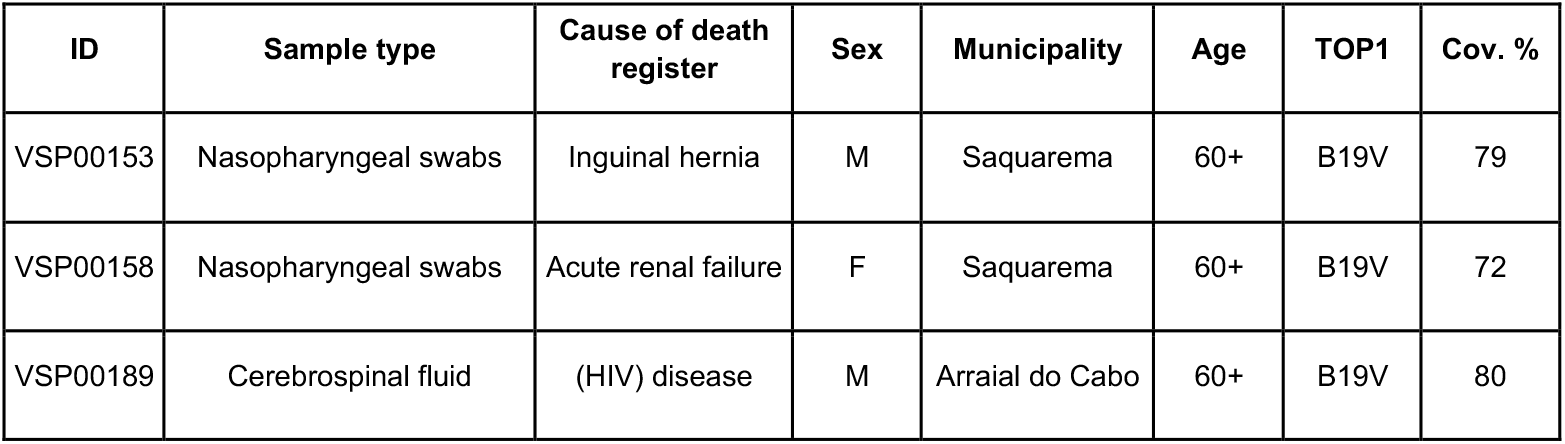
Fatal cases over 31 samples with B19V genomic coverage higher than 70%.

| ID | Sample type | Cause of death register | Sex | Municipality | Age | TOP1 | Cov. % |
| --- | --- | --- | --- | --- | --- | --- | --- |
| VSP00153 | Nasopharyngeal swabs | Inguinal hernia | M | Saquarema | 60+ | B19V | 79 |
| VSP00158 | Nasopharyngeal swabs | Acute renal failure | F | Saquarema | 60+ | B19V | 72 |
| VSP00189 | Cerebrospinal fluid | (HIV) disease | M | Arraial do Cabo | 60+ | B19V | 80 |

Phylogenetic analysis of genomic consensus sequences revealed that all identified B19V strains belonged to genotype 1A2 (Figure 3A). As NS1 is the primary marker for B19V, the six samples with 100% coverage of this gene were selected to construct a second phylogenetic tree, which confirmed genotype 1A2 (Figure 3B). Samples from Saquarema were positioned within the same clade. All samples were collected between April and May 2025, except for VSP00003, collected in January 2025 and more closely related to the reference for genotype 1A2 used.

**Figure 3:**
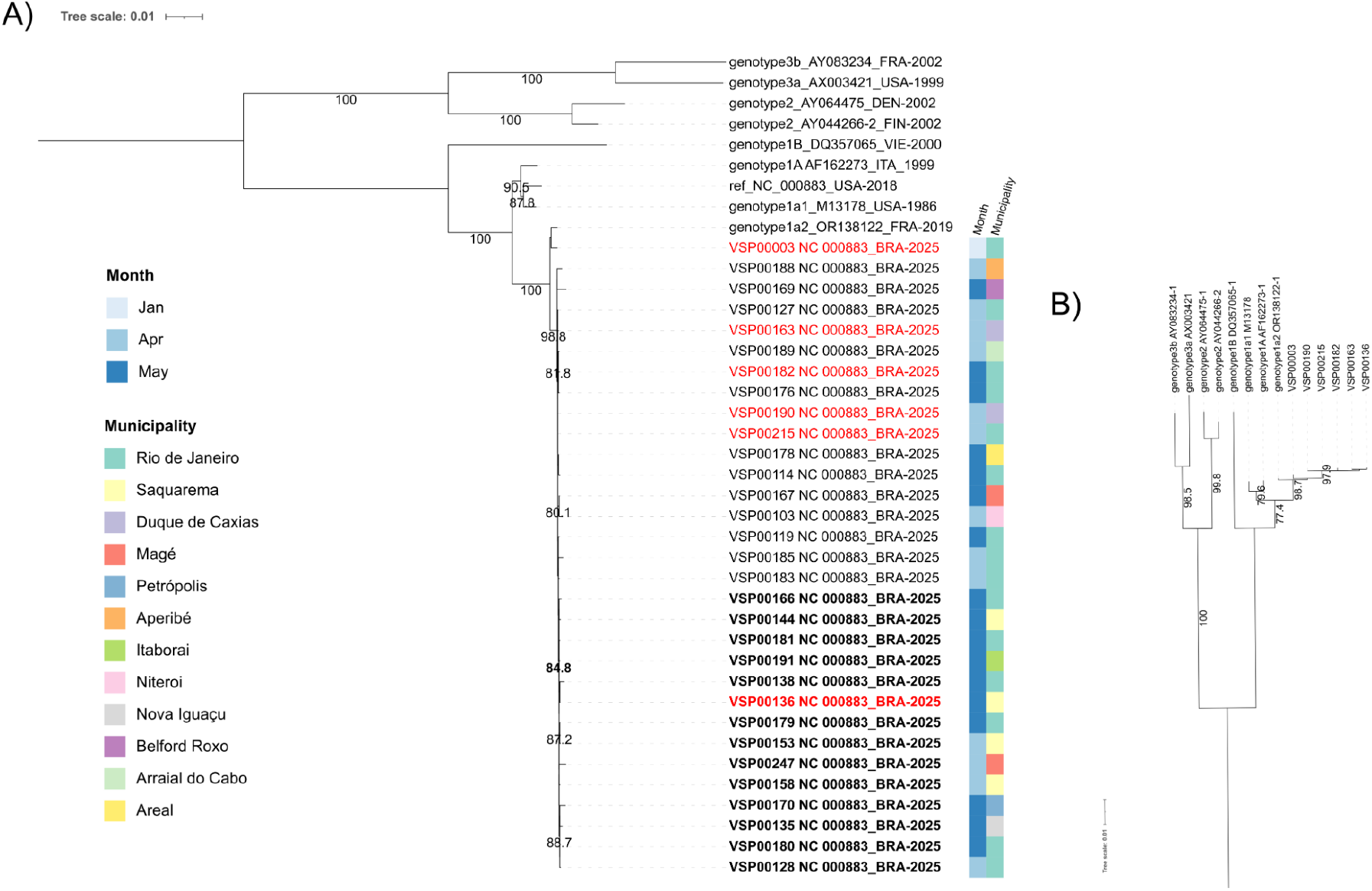
A) Consensus genome phylogenetic tree for 31 samples. B) NS1 gene phylogenetic tree for 6 samples. The substitution model for the maximum likelihood tree was chosen using ModelFinder [28]. Bootstrap values ≥ 70% were considered statistically significant and were reported. Trees were rooted by their midpoint and visualised in iTOL v7.5 (https://itol.embl.de/). Samples highlighted in red are those included in the NSP1 phylogenetic tree. Bold sample labels and bootstrap correspond to the Saquarema clade. BRA: Brazil; DEN: Denmark; FRA: France; ITA: Italy; FIN: Finland; VIE: Vietnam; USA: United States of America.

## Discussion

Recently, an increasing prevalence of B19V has been reported by European and North American researchers [3,8,12–15]. However, this pathogen remains neglected, and specific diagnostic tests are rarely performed in patients presenting atypical manifestations [4]. To our knowledge, contemporary Brazilian data on B19V circulation after the COVID-19 pandemic remain scarce [4]. The only post-pandemic collection data published, reported anti-B19V IgG seroprevalence among patients with hematological disorders from 2022 to 2023, which mainly reflects previous exposure rather than recent infection or active circulation [30]. Detection of B19V DNA, particularly in respiratory specimens, does not distinguish active infection from viral DNA persistence [2]. Nevertheless, several cases in our study exhibited high B19V genomic coverage in the absence of other detectable viral genomes. Our data shows a substantial circulation of B19V in severely ill and fatal cases and suggest a possible role for this pathogen in the clinical presentation observed in the patients.

The sequencing thresholds employed for the 31 B19V genomes highlighted in this study indicate high viral loads and true positive results [31]. Among this group, it is particularly noteworthy the number of elderly patients presenting exclusively B19V, most with fatal outcomes. Considering the higher seroprevalence for B19V reported in older people [30,32,33] and potential contribution of B19V to atypical extra-hematological presentations in adults [34], our observations warrants further investigation of the impact of B19V exposure into clinical manifestations in specific groups.

The main clinical indication among the patients of the present study was respiratory virus infections, but B19V was detected at a high frequency. Respiratory symptoms were observed in patients with B19V infection detected by quantitative PCR [3,35] and metagenomic approaches [16,36]. Notably, B19V was the unique virus detected with high coverage in patients presenting with meningitis and fatal renal failure in this study. Multiple neurological manifestations have been associated with B19V infection in the general population, with acute encephalitis occurring more frequently in children aged 0–5 years [1,34,37]. Here, cases of B19V-associated meningitis occurred in adults and infants, detected in CSF. Acute renal failure has been reported in association with B19V infection [34], but renal disease remains an uncommon clinical manifestation of this pathogen and typically progresses to spontaneous recovery in immunocompetent adults [4,38]. Therefore, although causality cannot be established, a potential contribution of B19V to the fatal outcome cannot be excluded, also considering the patient’s older age.

As the respiratory tract is a major route of B19V transmission and early replication, respiratory samples may be employed to support non-invasive surveillance [36,39]. Previous studies have demonstrated B19V detection in throat, oral and nasopharyngeal samples, highlighting the utility of unconventional specimens [16,36,40,41]. Consistently, hybrid capture sequencing detected B19V in this study, including several nasopharyngeal swabs, which cover severe and fatal cases without other viral pathogens identified at relevant coverage levels.

An important consideration is that the efficiency of capture sequencing critically depends on the quality and integrity of nucleic acids, particularly for degraded or low-abundance samples [18]. While thresholds for epidemiological and clinical studies for microorganisms employing hybrid capture approaches are not yet standardized, probe-based viral capture has shown that 10% genome coverage and 500 RPM can achieve sensitivity and specificity of 95% or higher [31]. Thus, the coverage and depth applied in this study are supported.

Molecular diagnosis of B19V infection is especially important for genotyping purposes. The phylogenetic classification of this virus comprises three distinct genotypes (1, 2, and 3). Genotype 1 is further divided into subtypes 1a and 1b, the former including groups 1a1 and 1a2. Group 1a2 tends to be more globally prevalent, with no clear functional advantage [42], being the presumed predominant genotype circulating in the European population in 2023 and 2024 [15]. In Brazil, genotype 1A is the most frequently reported, but other genotypes have also been described [7,33]. Our findings provide a more precise classification, particularly identifying the 1a2 group, which may contribute to improved monitoring.

B19V infection is not notifiable in many countries [4,12]. However, a pattern of 4-year cycles has been described in many countries, including Brazil [13,32,43]. Increasing post-pandemic reports, absence of a licensed vaccine, and limitation of current diagnostic methods may be associated with the high prevalence reported here [44–46]. In this study, just one case was previously investigated for B19V infection, with IgM serology reported as reactive. These observations suggest that B19V may represent an underrecognized and neglected viral infection, highlighting the need for improved surveillance, particularly during epidemic years and among high-risk groups, in whom B19V infection may lead to severe outcomes.

The present study revealed B19V detection across a broad range of ages, sample specimens, clinical presentations, and disease severities. The use of sequencing-based approaches to identify infections of unknown etiology, coinfections, and re-emerging pathogens, demonstrate the value of the genomic infrastructure expanded during the COVID-19 pandemic. Our findings contribute to strengthening B19V surveillance and reinforces the importance of considering this pathogen also in patients presenting atypical manifestations, especially during periods of increased viral circulation worldwide.

## Data Availability Statement

The sequences reported in this paper have been deposited in the GenBank database (accession nos. listed in Supplementary Table 1).

